# Dose-dependent smoking effects on depression inflammatory mediation and substantial prevention potential

**DOI:** 10.1101/2025.11.12.25339985

**Authors:** Zhenpeng Zhang, Tongyi Song, Xiaolei Wang, Jinwei Lu, Qiutong Yu, Zihan Wang, Aihua Yuan, Jinhua Sun, Bo Yuan

## Abstract

**Background:** Depressive disorders represent a leading cause of disability worldwide, yet prevention efforts have had limited success. Smoking is a modifiable risk factor frequently co-occurring with depression, but we lack quantitative evidence on dose-response relationships, cessation timelines, mediating biological pathways, and preventable disease burden.

**Methods:** We followed 169,741 UK Biobank participants free of baseline depression for 15 years (2006-2023), identifying 3,241 incident events. Using Cox models, we quantified associations between smoking phenotypes (status, intensity, pack-years, cessation timing) and depression incidence, mapped dose-response curves, and tested effect modification. We assessed inflammatory mediation, calculated population attributable fractions, and tested sensitivity to unmeasured confounding.

**Results:** Current smoking nearly doubled depression risk (HR 1.84, 95% CI 1.65-2.04), while previous smokers showed moderately elevated risk (HR 1.23, 95% CI 1.14-1.33). Cessation progressively reduced risk over subsequent decades. Dose-response was steep: 40 pack-years conferred 2.2-fold risk (HR 2.21, 95% CI 1.89-2.57), and 30 cigarettes daily doubled hazard (HR 2.16, 95% CI 1.71-2.72). Inflammatory biomarkers mediated 12.9% of the association (neutrophils HR 1.12, 95% CI 1.08-1.15; white blood cells HR 1.05, 95% CI 1.04-1.07), showing that most effects arise from non-inflammatory pathways. The population attributable fraction was 13.3%, representing one in eight preventable cases. Sensitivity analyses confirmed temporal stability and consistency across multiple analytical approaches; associations were unlikely to be explained by unmeasured confounding (E-value 3.08).

**Conclusions:** Smoking elevates depression risk in a dose-dependent manner, and cessation yields rapid, sustained reductions. Integrating tobacco cessation into mental health care could prevent one in eight depression cases. Inflammation mediates a minority of this effect; interventional studies are needed to test whether anti-inflammatory approaches provide additional benefit. Routine cessation services should be integrated into psychiatric and primary care settings.

## Background

Depressive disorders rank among the leading causes of disability worldwide, with a substantial portion of the global burden attributable to lifestyle and environmental factors[1–3]. Smoking co-occurs with depression in all demographic groups[4], and longitudinal cohorts increasingly show that tobacco exposure often precedes the onset of depressive symptoms[5]. This temporal sequence suggests smoking may be a modifiable risk factor rather than simply a comorbidity. Consistent with this view, large-scale Mendelian randomization studies have provided strong evidence that lifetime smoking is a causal risk factor for developing major depression[6, 7]. Clinicians still lack dose-specific guidance, clear estimates of how fast risk falls after quitting, and absolute risks and population-level burden to guide prevention.

Mood dysregulation can prompt smoking initiation and relapse. This two-way link makes effect estimates difficult to quantify and has slowed efforts to integrate tobacco control into psychiatric care[8]. The relationship is further complicated by shared genetic architecture; genome-wide association studies (GWAS) have identified substantial genetic correlations between nicotine dependence and depression. Recent work has begun to map how smoking patterns shape depression trajectories from starting age through persistence to relapse[9]. Many studies use a single exposure measure or stop follow-up at five to ten years[10]. Dose–response remains poorly described for cumulative pack-years and for time-dependent benefits after quitting[11]. Basic questions about conventional cigarettes are still open, while newer nicotine products such as e-cigarettes further complicate the picture and have themselves been linked to depression in large-scale surveys[12, 13].

Chronic inflammation may link tobacco toxins to mood dysregulation[14–16]. However, we do not know how much of the smoking effect operates through inflammation versus other pathways. Moreover, most studies report only relative hazards and omit population attributable fractions needed for policy prioritization[17], hampering resource allocation in mental health prevention.

Using 15 years of UK Biobank follow-up, we estimated how smoking status and intensity, cumulative pack-years, age at initiation, and cessation timing relate to depression incidence. Relative hazards were converted to absolute risk differences and population attributable fractions. We mapped dose–response and tested stability over time and across subgroups. Using biomarkers, we estimated the inflammatory component and used E-values to assess sensitivity to unmeasured confounding. These estimates are intended to guide prevention and support the integration of tobacco-cessation care into mental health services.

## Methods

### Study design and population

We used data from UK Biobank, a prospective cohort that recruited adults aged 40–69 years across England, Scotland, and Wales between 2006 and 2010[18]. The current data release includes 501,936 participants after excluding those who withdrew consent. The resource provides extensive phenotyping, biospecimens, and linkage to hospital and death records.

From this cohort, we applied sequential exclusion criteria (Figure 1A). First, we excluded participants with invalid follow-up (survival time ≤0 days or missing event/censoring dates), where survival time was calculated from baseline assessment to the earliest of depression diagnosis, death, loss to follow-up, or study end (31 December 2023). Second, we excluded participants with baseline depression or related mental health conditions, defined by any of the following five criteria: (1) mental health questionnaire status (Field 20126) indicating bipolar I/II disorder, probable recurrent major depression (severe or moderate), or single probable major depression episode; (2) ever feeling depressed for a whole week (Field 4598); (3) ever consulting a general practitioner for nerves, anxiety, or depression (Field 2090); (4) ever consulting a psychiatrist for nerves, anxiety, or depression (Field 2100); or (5) professional diagnosis (Field 20544) of depression, mania/hypomania/bipolar disorder, or social anxiety/phobia. Third, we excluded participants with missing smoking status. These exclusions yielded an analytical cohort of 280,816 eligible participants.

**Figure 1.**
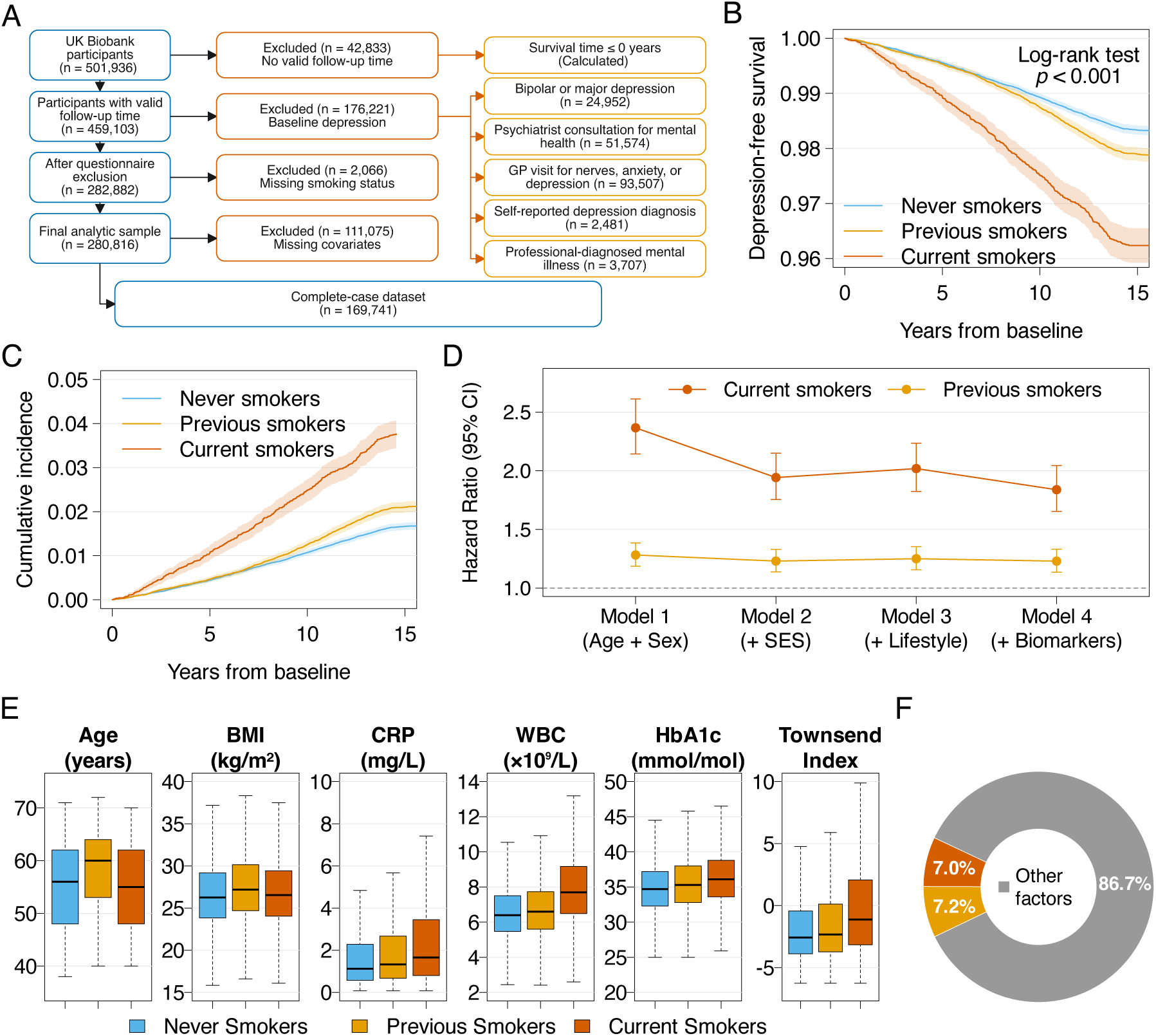
Smoking status and absolute burden of incident depression. **A:** Participant flow from 501,936 to 169,741 complete cases, with exclusions shown. **B:** Kaplan–Meier curves by smoking status (never, previous, current) with 95% CI bands; log-rank P<0.001. **C:** Cumulative incidence over 15 years by smoking status (never, previous, current), with shaded 95% confidence bands. Curves show event-free survival complement (1 minus survival probability). **D:** Forest plot showing hazard ratios across four nested models: Model 1 (age and sex only), Model 2 (+socioeconomic indicators), Model 3 (+lifestyle factors), Model 4 (+biomarkers). Estimates are shown separately for current and previous smokers versus never smokers. **E:** Box plots showing baseline distributions of age, body mass index (BMI), C-reactive protein (CRP), white blood cell count (WBC), glycated haemoglobin (HbA1c), and Townsend deprivation index by smoking status (never, previous, current). Boxes show medians and interquartile ranges (IQR); whiskers extend to 1.5×IQR. **F:** Population attributable fractions—current 7.0% (95% CI 5.8–8.3), previous 7.2% (6.0–8.5). Combined PAF 13.3% (11.5–15.2) from a joint multi-category model; category-specific PAFs are not additive. Estimated with Levin’s formula under full Model 4 adjustment.

For the primary analysis, we restricted to complete cases, defined as participants with non-missing data on all covariates: demographics (age, sex, Townsend deprivation index, household income, college education, employment status); lifestyle factors (body mass index, alcohol frequency, physical activity days, sleep duration); and biomarkers (C-reactive protein, white blood cell count, neutrophils, lymphocytes, monocytes, HbA1c, glucose, HDL cholesterol, LDL cholesterol, triglycerides, vitamin D, IGF-1, and sex hormone-binding globulin). We excluded five smoking dose variables (cigarettes per day, pack-years, age started smoking, age stopped smoking, and time to first cigarette) from the complete-case requirement due to high missingness (16.5%–94.2%). Complete-case analysis kept covariate sets identical across models.

### Exposure assessment

Baseline smoking status was self-reported as never, previous, or current. We obtained cigarettes per day, cumulative pack-years, ages at initiation and cessation, and time to first cigarette. Dose–response models excluded previous or current smokers with missing dose data; no imputation was performed for dose variables. All main-effects models used the same covariate set to ensure comparability. Years since quitting was calculated as baseline age minus age at cessation.

### Outcome assessment

Incident depression was ascertained from linked Hospital Episode Statistics records and death certificates using ICD-10 F32–F33. Participants with prior depression or probable depression at baseline were excluded. Follow-up time was calculated in years from baseline assessment to the earliest of depression diagnosis, death, loss to follow-up, or study end (31 December 2023). Median follow-up time and interquartile range (IQR) were computed across all participants in the complete-case cohort to characterize the duration of observation.

### Statistical analysis

Cox proportional hazards models were used to estimate associations with incident depression, with age included as a covariate. Four nested adjustment sets were used: (1) age and sex; (2) socioeconomic indicators; (3) lifestyle factors; (4) biomarkers. Dose–response for pack-years, cigarettes per day, and age at initiation was modeled with second-order polynomials within smoking-status strata. Cessation-duration models included previous smokers only.

Effect modification was examined for sex, age (<55/≥55 years), education, deprivation, and physical activity. Population attributable fractions for current and previous smoking were computed using Levin’s formula after full covariate adjustment [17].

Total, direct, and indirect effects were decomposed using a composite inflammation score based on neutrophils, monocytes, CRP, WBC, and lymphocytes. Mediation was quantified as the proportional reduction in the smoking log-hazard after adjusting for biomarkers versus the base model 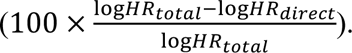 This coefficient-difference approach is descriptive and not a formal counterfactual mediation analysis; it assumes no uncontrolled exposure-mediator or mediator-outcome confounding and is interpreted as the share of the association explained by biomarker adjustment.

Analyses used R 4.5.1. Key packages and versions: arrow 21.0.0; colorspace 2.1.2; cowplot 1.2.0; data.table 1.17.8; ggplot2 4.0.0; grid 4.5.1; jsonlite 2.0.0; magick 2.9.0; magrittr 2.0.4; scales 1.4.0; splines 4.5.1; survival 3.8.3.

**Table 1.**
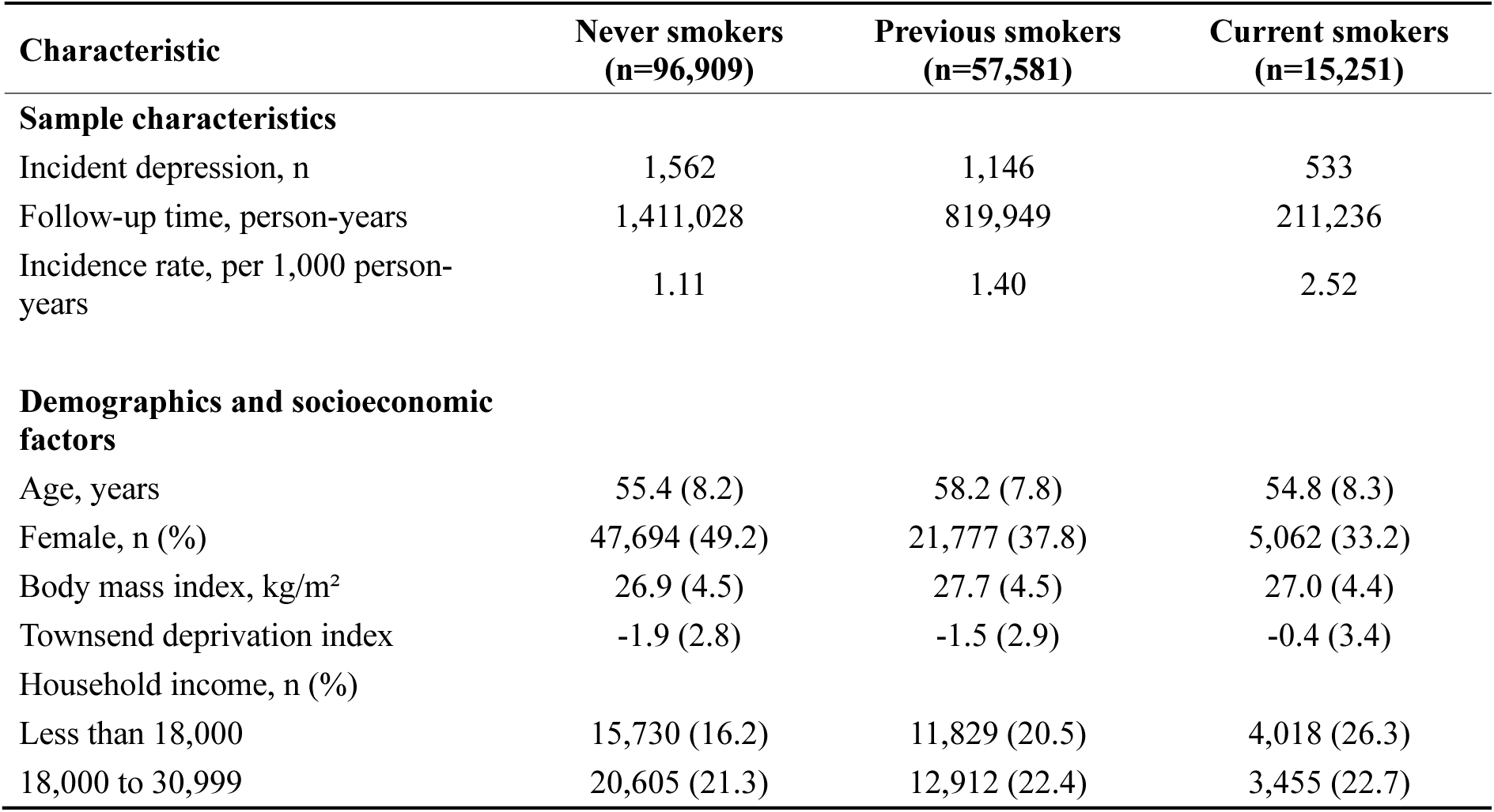
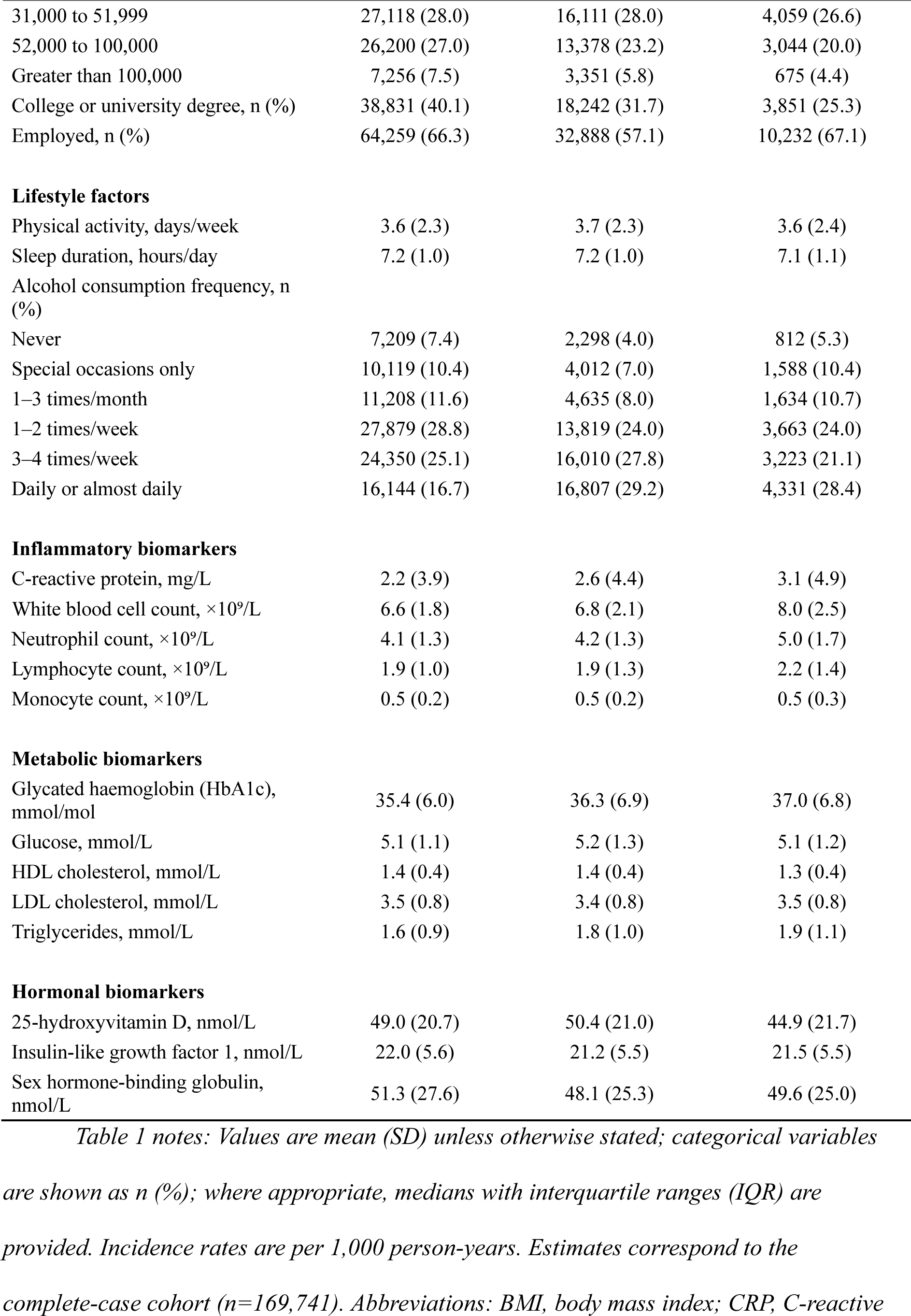
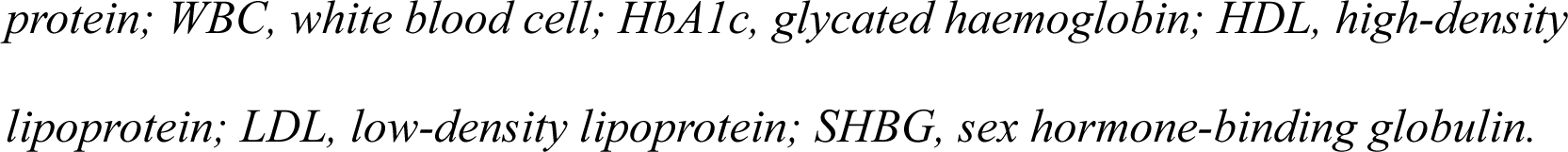
Baseline Characteristics.

notes: Values are mean (SD) unless otherwise stated; categorical variables are shown as n (%); where appropriate, medians with interquartile ranges (IQR) are provided.
| Characteristic | Never smokers<br>(n=96,909) | Previous smokers<br>(n=57,581) | Current smokers<br>(n=15,251) |
| --- | --- | --- | --- |
| **Sample characteristics** |  |  |  |
| Incident depression, n | 1,562 | 1,146 | 533 |
| Follow-up time, person-years | 1,411,028 | 819,949 | 211,236 |
| Incidence rate, per 1,000 person-years | 1.11 | 1.40 | 2.52 |
| **Demographics and socioeconomic factors** |  |  |  |
| Age, years | 55.4 (8.2) | 58.2 (7.8) | 54.8 (8.3) |
| Female, n (%) | 47,694 (49.2) | 21,777 (37.8) | 5,062 (33.2) |
| Body mass index, kg/m<sup>2</sup> | 26.9 (4.5) | 27.7 (4.5) | 27.0 (4.4) |
| Townsend deprivation index | -1.9 (2.8) | -1.5 (2.9) | -0.4 (3.4) |
| Household income, n (%) |  |  |  |
| Less than 18,000 | 15,730 (16.2) | 11,829 (20.5) | 4,018 (26.3) |
| 18,000 to 30,999 | 20,605 (21.3) | 12,912 (22.4) | 3,455 (22.7) |
| 31,000 to 51,999 | 27,118 (28.0) | 16,111 (28.0) | 4,059 (26.6) |
| 52,000 to 100,000 | 26,200 (27.0) | 13,378 (23.2) | 3,044 (20.0) |
| Greater than 100,000 | 7,256 (7.5) | 3,351 (5.8) | 675 (4.4) |
| College or university degree, n (%) | 38,831 (40.1) | 18,242 (31.7) | 3,851 (25.3) |
| Employed, n (%) | 64,259 (66.3) | 32,888 (57.1) | 10,232 (67.1) |
| **Lifestyle factors** |  |  |  |
| Physical activity, days/week | 3.6 (2.3) | 3.7 (2.3) | 3.6 (2.4) |
| Sleep duration, hours/day | 7.2 (1.0) | 7.2 (1.0) | 7.1 (1.1) |
| Alcohol consumption frequency, n (%) |  |  |  |
| Never | 7,209 (7.4) | 2,298 (4.0) | 812 (5.3) |
| Special occasions only | 10,119 (10.4) | 4,012 (7.0) | 1,588 (10.4) |
| 1–3 times/month | 11,208 (11.6) | 4,635 (8.0) | 1,634 (10.7) |
| 1–2 times/week | 27,879 (28.8) | 13,819 (24.0) | 3,663 (24.0) |
| 3–4 times/week | 24,350 (25.1) | 16,010 (27.8) | 3,223 (21.1) |
| Daily or almost daily | 16,144 (16.7) | 16,807 (29.2) | 4,331 (28.4) |
| **Inflammatory biomarkers** |  |  |  |
| C-reactive protein, mg/L | 2.2 (3.9) | 2.6 (4.4) | 3.1 (4.9) |
| White blood cell count, $\times 10^9/L$ | 6.6 (1.8) | 6.8 (2.1) | 8.0 (2.5) |
| Neutrophil count, $\times 10^9/L$ | 4.1 (1.3) | 4.2 (1.3) | 5.0 (1.7) |
| Lymphocyte count, $\times 10^9/L$ | 1.9 (1.0) | 1.9 (1.3) | 2.2 (1.4) |
| Monocyte count, $\times 10^9/L$ | 0.5 (0.2) | 0.5 (0.2) | 0.5 (0.3) |
| **Metabolic biomarkers** |  |  |  |
| Glycated haemoglobin (HbA1c), mmol/mol | 35.4 (6.0) | 36.3 (6.9) | 37.0 (6.8) |
| Glucose, mmol/L | 5.1 (1.1) | 5.2 (1.3) | 5.1 (1.2) |
| HDL cholesterol, mmol/L | 1.4 (0.4) | 1.4 (0.4) | 1.3 (0.4) |
| LDL cholesterol, mmol/L | 3.5 (0.8) | 3.4 (0.8) | 3.5 (0.8) |
| Triglycerides, mmol/L | 1.6 (0.9) | 1.8 (1.0) | 1.9 (1.1) |
| **Hormonal biomarkers** |  |  |  |
| 25-hydroxyvitamin D, nmol/L | 49.0 (20.7) | 50.4 (21.0) | 44.9 (21.7) |
| Insulin-like growth factor 1, nmol/L | 22.0 (5.6) | 21.2 (5.5) | 21.5 (5.5) |
| Sex hormone-binding globulin, nmol/L | 51.3 (27.6) | 48.1 (25.3) | 49.6 (25.0) |

*protein; WBC, white blood cell; HbA1c, glycated haemoglobin; HDL, high-density lipoprotein; LDL, low-density lipoprotein; SHBG, sex hormone-binding globulin.*

With 3,241 events in 169,741 participants, power to detect a hazard ratio of 1.20 for main effects exceeded 99% at α=0.05 (two-sided). For subgroup analyses, strata ranged from approximately 2,000 to 50,000 participants with 50 to 1,200 events, providing adequate power for moderate to large interactions.

### Sensitivity analyses

To assess the stability of our findings, we conducted three sensitivity analyses addressing key threats to validity.

We evaluated the temporal stability of associations by splitting the follow-up period into five intervals (0–3, 3–6, 6–9, 9–12, and ≥12 years from baseline) and estimating separate hazard ratios within each window. This analysis addresses concerns about reverse causation in early follow-up and tests for time-varying effects over extended observation periods.

We compared estimates across three analytical approaches to test sensitivity to adjustment choices and potential selection bias: (1) Model 4 (primary complete-case analysis, n=169,741) with full adjustment including all demographic, socioeconomic, lifestyle, and biomarker covariates; (2) Model 3 equivalent (n=169,741), adjusting for demographic, socioeconomic, and lifestyle factors but excluding biomarkers to assess potential over-adjustment bias; and (3) a partial missing indicator method (n=195,651, 69.7% of eligible) that added binary missing indicators and median imputation for six biomarkers with high missingness (glucose, HDL, SHBG, vitamin D, LDL, and triglycerides) while still requiring complete data on other covariates. The third approach addresses potential selection bias from complete-case analysis, which excluded 39.6% of eligible participants due to missing covariate data, by testing whether participants with biomarker missingness differ systematically in the smoking-depression association. Consistency across these three models would indicate that results are not driven by specific adjustment decisions or by the subset of participants with complete biomarker data.

We quantified the resistance of associations to unmeasured confounding by calculating E-values for both the point estimate and the lower 95% confidence interval bound. The E-value represents the minimum strength of association that an unmeasured confounder would need with both the exposure (smoking) and outcome (depression) to fully explain away the observed association, conditional on measured covariates. E-values were computed as 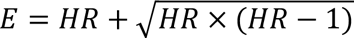 for hazard ratios above 1, following the approach of VanderWeele and Ding. Higher E-values indicate greater resistance to potential unmeasured confounding.

All sensitivity analyses used the same outcome definition (incident depression from ICD-10 codes F32–F33 in hospital records or death certificates) and primary exposure definition (baseline smoking status) as the main analysis. Results were considered stable if effect estimates remained materially unchanged across sensitivity analyses, accounting for statistical uncertainty through confidence intervals.

## Results

Among 280,816 eligible adults, 169,741 (60.4%) had complete data (Figure 1A). The full eligible cohort included 5,852 incident depression events. The complete-case restriction excluded 111,075 participants (39.6%) with incomplete covariate data, among whom 2,611 (calculated: 5,852-3,241) events occurred, leaving 3,241 events in the analytical sample. Over a median 15.0 years of follow-up (IQR 14.2–15.6; range 0.0–17.0 years; mean 14.4±2.4 years), these 3,241 incident depression diagnoses accumulated 2,442,213 person-years of observation (Table 1). Excluded participants had higher baseline CRP (2.65 vs 2.40 mg/L) and lived in more deprived areas (Townsend -1.33 vs -1.62), suggesting our estimates are likely conservative.

Table 1 presents baseline characteristics of the 169,741 participants stratified by smoking status, including demographics, socioeconomic factors, lifestyle behaviours, and biomarkers. In the complete-case cohort, crude incidence per 1,000 person-years was 1.11 (95% CI 1.05–1.17) for never smokers, 1.40 (1.32–1.49) for previous smokers, and 2.52 (2.31–2.75) for current smokers.

Kaplan-Meier curves showed clear separation between smoking groups throughout the follow-up period (Figure 1B). Current smokers exhibited the lowest event-free survival, followed by previous smokers, while never smokers maintained the highest survival probability. The three curves diverged early and stayed distinct across the entire observation period, with log-rank test confirming highly significant differences (P<0.001).

Ten-year cumulative incidence was 1.07% for never, 1.25% for previous, and 2.49% for current smokers (Figure 1C). After full adjustment, hazards were higher for previous (HR 1.23, 95% CI 1.14–1.33) and current smokers (HR 1.84, 95% CI 1.65–2.04) versus never smokers (Figure 1D). There were 533 events among 15,251 current smokers (Table 1) and 1,562 among 96,909 never smokers (Table 1). The absolute excess was 1.41 per 1,000 person-years (calculated from Table 1). Sequential adjustment moved the current-smoking HR from 2.37 (Model 1) to 1.84 (Model 4) (Figure 1D), suggesting that socioeconomic, lifestyle, and biomarker factors explain approximately 29% of the excess log-hazard.

Baseline characteristics differed across smoking groups (Figure 1E). Never smokers were slightly younger than previous smokers but older than current smokers (means 55.4±8.2, 58.2±7.8, 54.8±8.3 years; medians 56, 60, 55 years). Previous smokers had the highest BMI (mean 27.7±4.5 kg/m², median 27.2), followed by current (27.0±4.4, median 26.5) and never smokers (26.9±4.5, median 26.2). Inflammatory markers increased with smoking exposure: CRP means were 2.2±3.9, 2.6±4.4, and 3.1±4.9 mg/L for never, previous, and current smokers (medians 1.12, 1.33, 1.66), while WBC counts were 6.6±1.8, 6.8±2.1, and 8.0±2.5 ×10⁹/L (medians 6.4, 6.6, 7.7). Current smokers had higher HbA1c (mean 37.0±6.8 mmol/mol, median 36.1) than previous (36.3±6.9, median 35.3) and never smokers (35.4±6.0, median 34.7). Socioeconomic gradients were pronounced: current smokers lived in more deprived areas (Townsend mean −0.4±3.4, median −1.12) compared to previous (−1.5±2.9, median −2.33) and never smokers (−1.9±2.8, median −2.58). Women made up 49.2% of never smokers and 33.2% of current smokers (Table 1).

Population attributable fractions (PAFs) totaled 13.3% (combined, from a joint multi-category model; category-specific PAFs—current 7.0%, previous 7.2%—are not additive) (Figure 1F), corresponding to ≈430 preventable cases over 15 years.

These main effects confirm that smoking elevates depression risk. To provide dose-specific estimates, we examined dose-response curves across four smoking metrics. All four metrics showed steep, nonlinear relationships with depression incidence (P for trend < 0.001; Figure 2). Both unadjusted (solid lines) and adjusted (dashed lines, Model 4) curves followed similar nonlinear patterns.

**Figure 2.**
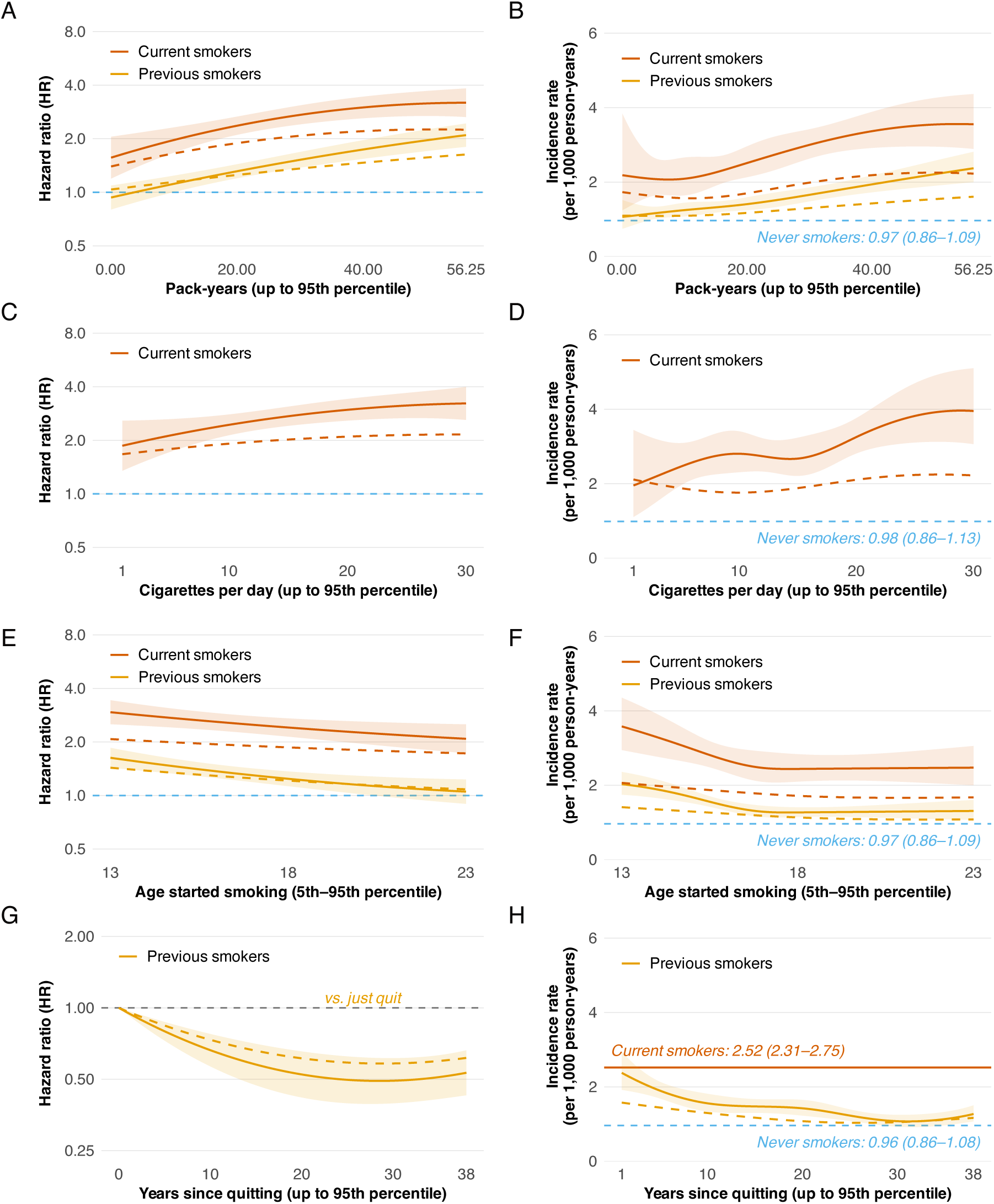
Dose–response between smoking intensity and depression risk. **A:** Pack-years plotted against hazard ratio (HR). Solid lines show unadjusted estimates; dashed lines show fully adjusted estimates (Model 4) for current and previous smokers. **B:** Pack-years plotted against incidence rate (per 1,000 person-years). Solid lines show unadjusted estimates; dashed lines show fully adjusted estimates (Model 4) for current and previous smokers. **C:** Daily cigarette consumption plotted against hazard ratio (HR). Solid lines show unadjusted estimates; dashed lines show fully adjusted estimates (Model 4) for current and previous smokers. **D:** Daily cigarette consumption plotted against incidence rate (per 1,000 person-years). Solid lines show unadjusted estimates; dashed lines show fully adjusted estimates (Model 4) for current and previous smokers. **E:** Age at smoking initiation plotted against hazard ratio (HR). Solid lines show unadjusted estimates; dashed lines show fully adjusted estimates (Model 4) for current and previous smokers. **F:** Age at smoking initiation plotted against incidence rate (per 1,000 person-years). Solid lines show unadjusted estimates; dashed lines show fully adjusted estimates (Model 4) for current and previous smokers. **G:** Years since quitting smoking plotted against hazard ratio (HR) in previous smokers. Solid lines show unadjusted estimates; dashed lines show fully adjusted estimates (Model 4). **H:** Years since quitting smoking plotted against incidence rate (per 1,000 person-years) in previous smokers. Solid lines show unadjusted estimates; dashed lines show fully adjusted estimates (Model 4).

Pack-years showed steep dose-response curves (Figure 2A-B). Both hazard ratios (Figure 2A) and incidence rates (Figure 2B) increased nonlinearly with cumulative exposure. In current smokers, fully adjusted HRs rose from 1.39 at 0 pack-years to 1.89 at 20 and 2.21 at 40 (Figure 2A), with the steepest gradient in the first 20 pack-years (≈0.025 per pack-year). Unadjusted estimates showed similar patterns but higher absolute values. Incidence rates paralleled these trends (Figure 2B), rising from approximately 2.0 to over 3.5 per 1,000 person-years at 40 pack-years in current smokers. Previous smokers exhibited gentler slopes in both metrics (HR 1.25 at 20 pack-years; 1.47 at 40; P for trend < 0.001), averaging ≈0.011 per pack-year, approximately half the rate observed in current smokers.

Daily cigarette consumption displayed rising curves with plateaus near 30 cigarettes per day in both HR (Figure 2C) and incidence rate (Figure 2D). At this level, fully adjusted HR reached 2.16 (95% CI 1.71–2.72), while incidence rates approached 3.0 per 1,000 person-years, suggesting diminishing marginal increases in risk at very high consumption levels.

Age at smoking initiation showed inverse associations in both HR (Figure 2E) and incidence rate (Figure 2F). Starting smoking at younger ages (e.g., 13 years) conferred higher risk (fully adjusted HR ≈2.1 for current smokers, incidence rate ≈2.5 per 1,000 person-years) compared to later initiation (e.g., 25 years: HR ≈1.5, incidence rate ≈1.8 per 1,000 person-years). This gradient was evident in both metrics, with earlier initiation consistently predicting worse outcomes in current and previous smokers.

Years since quitting showed rapid early benefits in both HR (Figure 2G) and incidence rate (Figure 2H). Compared with recent quitters, fully adjusted HR declined to 0.96 at 1 year, 0.84 at 5 years, 0.73 at 10 years, and 0.61 at 20 years (Figure 2G), representing risk reductions of 4%, 16%, 27%, and 39% respectively. Incidence rates showed corresponding temporal declines (Figure 2H), progressively approaching never-smoker levels. Both unadjusted and adjusted estimates revealed that most benefits accrued within the first decade after cessation.

At the same cumulative exposure (e.g., 40 pack-years), current smoking carried substantially more risk than past smoking (HRs 2.21 vs 1.47; ∼50% higher), highlighting the benefits of quitting beyond simple dose reduction.

These dose-response patterns represent population averages. To identify which subgroups face disproportionate risk, we examined effect modification by demographics, socioeconomic status, and lifestyle factors.

Smoking effects varied minimally by sex (Pinteraction=0.259), age (0.663), education (0.976), or deprivation (0.294), with one exception: physical activity (Pinteraction=0.035; Figure 3). The forest plot displays hazard ratios for current versus never smokers (vermilion diamonds) and previous versus never smokers (orange diamonds) across five subgroup variables. Each estimate includes 95% confidence intervals, sample sizes, and event counts. Among current smokers, the physical activity interaction showed a complex pattern: HRs were 1.41 with no activity, 2.48 with low activity (1–3 days/week), 1.51 with moderate activity (4–5 days/week), and 1.93 with high activity (≥6 days/week), with low activity showing unexpectedly elevated risk; estimates were less precise in smaller physical activity strata. Although current-smoking women showed numerically higher HRs than men (2.08 vs 1.70) and a deprivation gradient was observed (HR 1.97 in the most vs 1.70 in the least deprived tertile), these differences did not reach statistical significance (Pinteraction=0.259 and 0.294, respectively).

**Figure 3.**
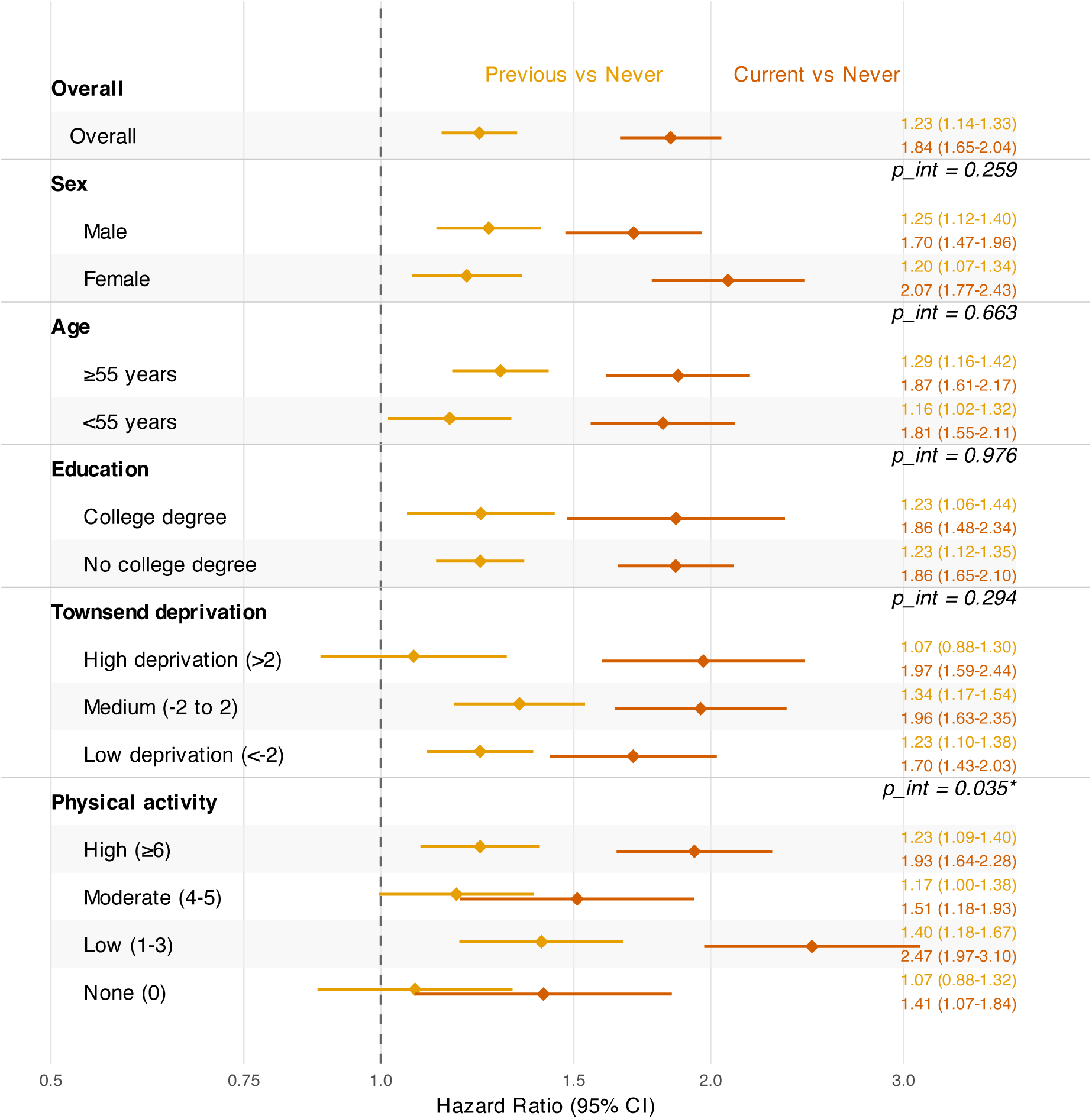
Subgroup analysis of current and previous smoking versus never smoking, by sex, age, education, deprivation, and physical activity. Forest plot showing hazard ratios for current versus never smoking (vermilion diamonds) and previous versus never smoking (orange diamonds) across five subgroup variables. Each estimate includes 95% confidence intervals, sample sizes, and event counts. Error bars represent 95% confidence intervals. Tests for interaction assess whether smoking effects differ across subgroups. Reference line at HR = 1.0.

Extended subgroup analyses across socioeconomic factors (Figure S1), lifestyle behaviours (Figure S2), inflammatory biomarkers (Figure S3), metabolic markers (Figure S4), and hormonal indicators (Figure S5) revealed consistent smoking-depression associations across most strata, showing that the link holds across diverse biological profiles.

To design better interventions, we need to understand the mechanisms. We therefore quantified inflammatory mediation to determine whether this pathway substantially contributes to the smoking-depression association.

We quantified inflammatory mediation in four steps. First, we confirmed that smoking elevates inflammatory biomarkers (Figure 4A). Current smokers showed the highest mean levels for CRP (3.1±4.9 mg/L), WBC (8.0±2.5 ×10⁹/L), neutrophils (5.0±1.7 ×10⁹/L), and monocytes (0.5±0.3 ×10⁹/L), followed by previous smokers and never smokers, showing a clear gradient of systemic inflammation associated with smoking exposure.

**Figure 4.**
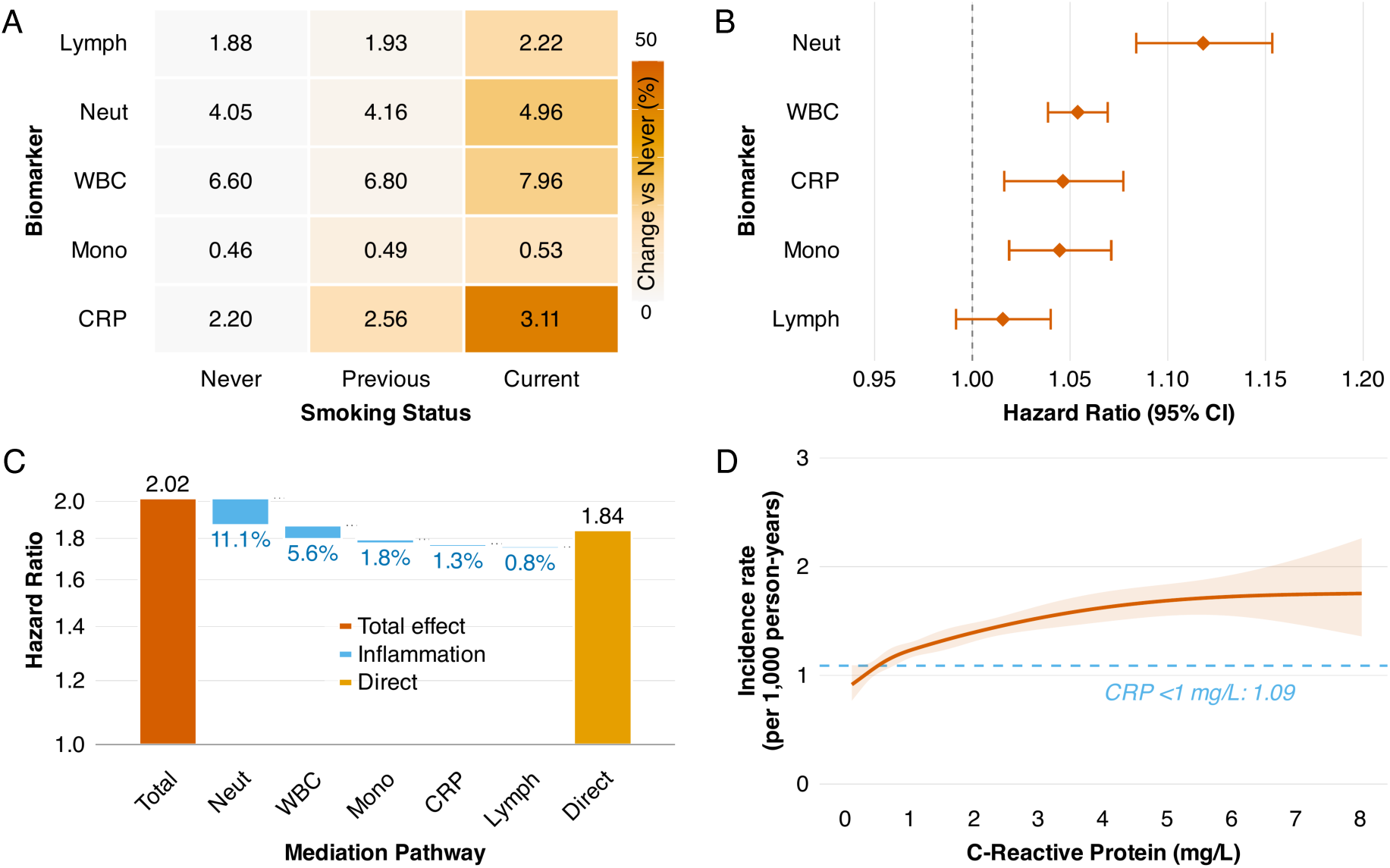
Biomarkers in the smoking–depression pathway. **A:** Heatmap showing mean levels of inflammatory biomarkers (CRP, WBC, neutrophils, lymphocytes, monocytes) by smoking status (never, previous, current). Cell values show mean biomarker levels. Color intensity represents relative change from never smokers (%). **B:** Forest plot showing hazard ratios for depression per standard deviation increase in each inflammatory biomarker, adjusted for smoking status and all Model 4 covariates. Error bars represent 95% confidence intervals. Reference line at HR = 1.0. **C:** Waterfall plot showing mediation analysis results. Bars represent the proportion of the total smoking-depression effect explained by each inflammatory pathway. Total effect HR = 2.02 (current vs never), direct effect HR = 1.84 after adjustment for inflammatory markers. Individual biomarker contributions: neutrophils 11.1%, WBC 5.6%, monocytes 1.8%, CRP 1.3%. Combined inflammatory mediation = 12.9%. **D:** Dose-response relationship between baseline CRP and depression incidence. Red curve shows crude incidence rate per 1,000 person-years across CRP levels, fitted using Poisson regression with natural splines (df=4). Shaded area represents 95% confidence interval. Blue dashed line indicates reference rate for CRP <1 mg/L. Demonstrates increasing depression risk with higher systemic inflammation.

After adjusting for smoking status, neutrophils showed the strongest association with depression (HR 1.12 per SD increase), followed by WBC (1.05), CRP (1.05), and monocytes (1.04); lymphocytes showed no association (HR 1.02, 95% CI 0.99–1.04) (Figure 4B).

Neutrophils explained the largest share of mediation (11.1%), followed by WBC (5.6%), monocytes (1.8%), and CRP (1.3%); collectively, inflammatory markers accounted for 12.9% of the total current smoking effect, with a remaining direct effect of HR 1.84 (total effect HR 2.02) (Figure 4C). This pattern points to innate immune activation. Because these markers are correlated, the summed shares exceed 12.9%.

Higher baseline CRP levels were associated with increased depression incidence in a dose-dependent manner (Figure 4D). Using natural spline regression, the crude incidence rate rose from approximately 1.3 per 1,000 person-years at CRP <1 mg/L to over 2.0 per 1,000 person-years at CRP ≥5 mg/L, showing a clear gradient consistent with a role for systemic inflammation in depression pathogenesis.

Sensitivity analyses confirmed consistency across three dimensions (Figure 5). Missing data patterns varied by covariate type (Figure 5A). Core variables (age, sex, smoking status) had complete data. Sociodemographic variables showed modest missingness: household income (15.5%), physical activity (5.0%), education (2.0%). Inflammatory markers had low to moderate missingness: CRP (6.5%), WBC and differential counts (4.7–4.8%). Metabolic and hormonal biomarkers showed higher rates: SHBG (14.8%), glucose and HDL (14.1%), vitamin D (10.3%), and lipids (6.4–6.9%). The complete-case analysis excluded 39.6% of participants due to any missing covariate.

**Figure 5.**
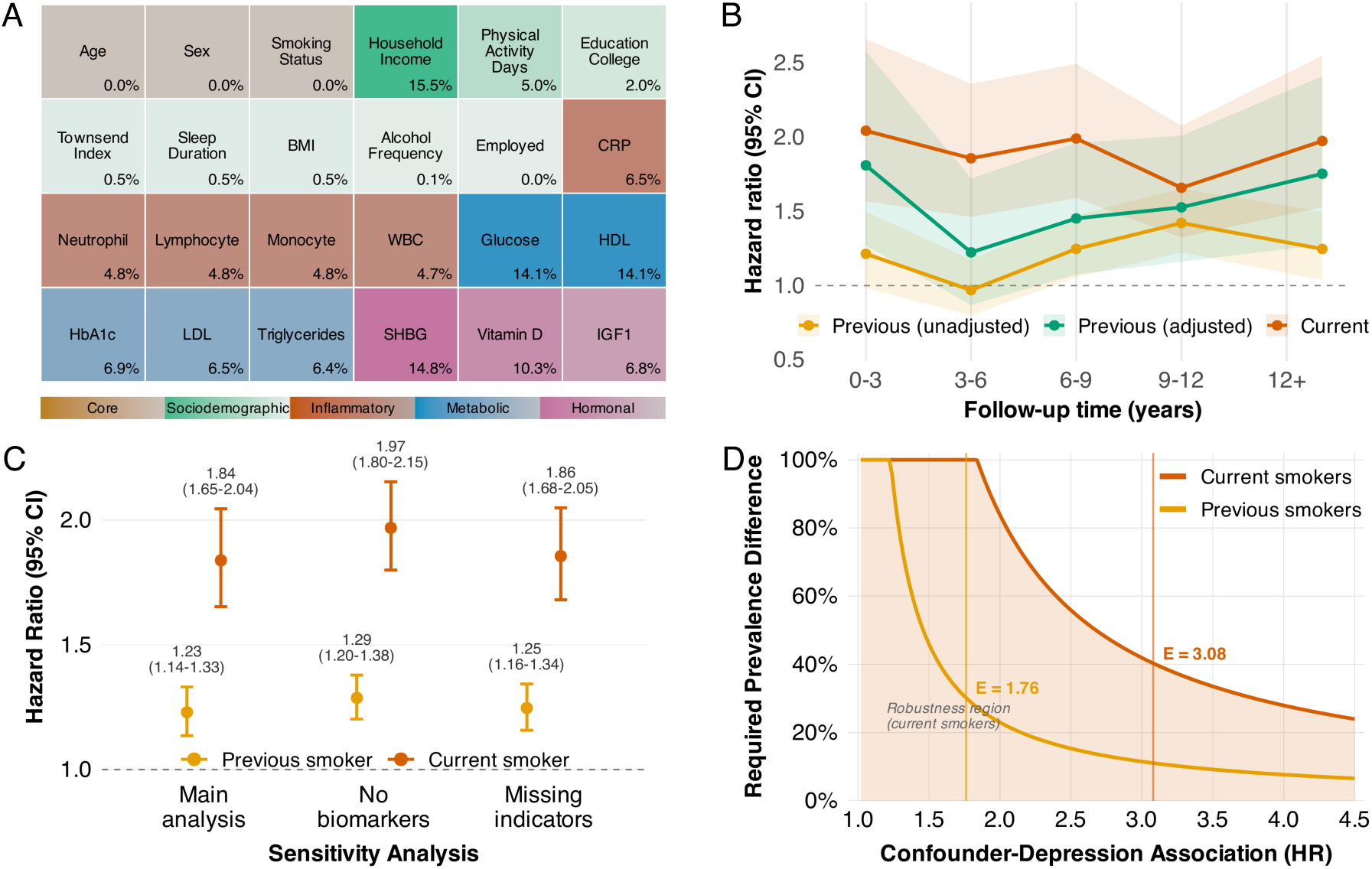
Sensitivity analyses. **A:** Heatmap showing percentage of missing data for each covariate, grouped by variable type (Core, Sociodemographic, Inflammatory, Metabolic, Hormonal). Color intensity represents missing percentage. Used to assess potential selection bias in complete-case analysis (n=169,741) versus full eligible cohort (n=280,816). **B:** Time-stratified hazard ratios for current and previous versus never smoking across five follow-up windows: 0–3, 3–6, 6–9, 9–12, and ≥12 years since baseline. Error bars represent 95% confidence intervals. Demonstrates temporal stability of associations and tests proportional hazards assumption. **C:** Forest plot comparing hazard ratios for current and previous versus never smoking across three analytical approaches: (1) Main analysis (Model 4, fully adjusted with all biomarkers, n=169,741), (2) Without biomarkers (Model 3 equivalent), (3) Partial missing indicators method (n=195,651, 69.7% of eligible participants). Error bars represent 95% confidence intervals. Assesses consistency with different adjustment strategies and missing data handling. **D:** E-value curves. For current versus never smoking (Model 4), E-value = 3.08 for the point estimate (HR 1.84) and 2.69 for the lower 95% CI bound (HR 1.65), computed as 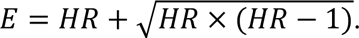

Hazard ratios stayed elevated across five time windows (Figure 5B). For current smokers, HRs ranged from 2.04 in the earliest window (0–3 years) to 1.97 in the latest window (≥12 years), with each window containing >90 events supporting stable estimates. Former smokers also showed consistently higher risk across all windows.

Results were consistent across the three analytical approaches described in Methods (Figure 5C). For current smoking, HRs were similar in the primary complete-case analysis, the model excluding biomarkers, and the missing indicators analysis. Former smokers showed similarly consistent estimates across all three approaches, supporting the findings regardless of adjustment choices and potential selection bias from missing data.

E-values indicated strong resilience to unmeasured confounding. For current versus never smoking, the E-value was 3.08 for the point estimate and 2.69 for the lower 95% CI bound, suggesting that unmeasured confounding would need to be very strong to nullify the observed association (Figure 5D).

## Discussion

Our findings establish smoking as a strong, dose-dependent driver of incident depression, not just a correlate. Mendelian randomization studies support this causal interpretation[6]. Quitting reduces risk: meta-analyses show post-cessation improvements in depressive and anxiety symptoms that match the effect sizes of antidepressant treatment[19], and pharmacotherapies such as varenicline, cytisine, and combination nicotine replacement therapy increase long-term abstinence rates[20]. Inflammation explains only a minority (12.9%) of the excess risk, suggesting that non-inflammatory neurobiological pathways drive most of the association. We also found that smoking effects varied by physical activity level, suggesting that behavioural interventions should bundle cessation support with physical activity promotion rather than treating them separately.

In this cohort with 2.4 million person-years of follow-up, current smoking nearly doubled the hazard of incident depression, with risk remaining elevated even decades after quitting. The associations show steep dose–response patterns across pack-years, daily consumption, age at initiation, and years since quitting. At the population level, smoking accounts for 13.3% of incident depression, representing approximately one in eight cases. The absolute excess of 1.41 cases per 1,000 person-years translates to roughly one extra diagnosis for every 700 smokers followed for a year. Risk declines within the first year after quitting and continues to fall through the first decade, with benefits persisting for at least two decades.

Most of the effect likely arises from non-inflammatory pathways. These include neurochemical, neuroendocrine, and vascular mechanisms that should be addressed alongside inflammation in future interventions. Chronic nicotine exposure alters nicotinic acetylcholine receptor (nAChR) density and function[21]. These adaptations dysregulate dopamine, serotonin, and other neurotransmitter systems[22], disrupting mood regulation. Recent evidence shows that nicotine engages a VTA-NAc feedback loop that inhibits amygdala-projecting dopamine neurons, inducing anxiety-like behaviours[23]. During withdrawal, these disrupted circuits may exacerbate negative affect and increase depression risk even after cessation.

These findings extend prior work[8, 9]. We quantified several smoking patterns in parallel, converted relative hazards into absolute burden, and estimated the share explained by inflammation. Results were consistent with international cohorts[5]. Heavy smokers and socioeconomically disadvantaged groups bore disproportionate risk, marking them as priority targets for integrated cessation support. We also found an interaction with physical activity[10]. Sedentary smokers faced a double hit, so behavioural care should bundle cessation with increasing activity rather than treat them separately. Women and residents of high-deprivation areas faced higher risks, calling for equity-focused, culturally tailored cessation programmes that include mental-health screening and support. One-size-fits-all approaches often underserve the people who carry the heaviest combined burden of smoking and mood disorders.

This mirrors cardiovascular and cancer benefit timelines[24] and provides practical guidance for motivational interviewing: tell patients that quitting today can lower depression risk within a year, with gains that build over time. The mortality benefits of cessation are also profound and appear within a few years of quitting, reinforcing the urgency of intervention[25]. Even so, risk stays slightly above never-smoker levels two decades after cessation; previous smokers still need mental-health follow-up and relapse prevention.

Inflammatory mediation was 12.9%, but it still carries clinical and mechanistic value[14, 26]. This supports pairing cessation with adjunct anti-inflammatory interventions, particularly when baseline inflammatory markers are elevated. These include exercise, a Mediterranean diet, or targeted pharmacological agents. However, the role of specific inflammatory markers is complex. Mendelian randomization analyses suggest that while inflammatory pathways are implicated in depression, the comorbidity with coronary heart disease seems to arise from shared environmental factors rather than shared genetics, though IL-6 and CRP are still highlighted as likely causal risk factors for depression itself[27]. Our finding that neutrophils are a stronger mediator than CRP refines this understanding.

CRP already functions as a clinical warning signal for heightened cardiovascular disease risk, and recent syntheses emphasise how psychiatric disorders and cardiovascular outcomes are closely linked[28]. Inflammatory biomarkers, particularly CRP and inflammatory cell counts, represent shared risk factors linking psychiatric disorders to cardiovascular sequelae[15, 16]. Observing the same marker in our mediation pathway suggests that smoking-driven inflammation can amplify both depression and downstream cardiometabolic burden, making cessation a two-pronged prevention strategy. Observational data from the UK Biobank likewise show that common psychiatric disorders substantially elevate subsequent cardiovascular hospitalizations regardless of inherited susceptibility, highlighting the shared downstream burden once mood disorders take hold[29].

Neutrophils mattered more than lymphocytes, implicating innate rather than adaptive immunity; interventional studies are required to test whether targeting innate-immune pathways improves mental-health outcomes beyond standard cessation care.

From a policy perspective, a 13.3% population attributable fraction makes smoking cessation a high-impact prevention strategy for depression, comparable to many psychosocial interventions prioritized in current mental health policy. Countries with advanced tobacco control measures and low smoking prevalence are increasingly adopting tobacco endgame strategies[30], which could also improve mental health while reducing physical disease burden.

Integrating cessation support into psychiatric and primary care workflows[31] through universal screening, motivational counselling, pharmacotherapy (varenicline, bupropion, nicotine replacement), and relapse monitoring could prevent approximately 430 depression cases (calculated: 3,241 × 0.133) per 169,741 mid-life adults over 15 years. Benefits also accrue to cardiometabolic health, quality of life, and healthcare costs. These gains require sustained investment. Brief, sporadic advice changes little; intensive, multi-contact programmes achieve 20–30% long-term abstinence.

Hazard ratios were elevated beyond 12 years. The high E-value (3.08) makes it unlikely that unmeasured confounding fully explains the results. The evidence supports investment in integrated tobacco–mental-health programmes.

We note several limitations. Smoking status was self-reported and may be misclassified. Even so, biochemical validation studies suggest high specificity for current smoking.

Complete-case analysis excluded 39.6% of eligible participants due to missing covariate data. Among the 28 analysis variables, missingness was highest for household income (15.5%), sex hormone-binding globulin (14.8%), and glucose (14.1%). Compared with excluded individuals, the complete-case sample was younger (mean difference: -1.3 years), had 10.3% fewer women, and showed lower inflammatory markers (C-reactive protein: 2.40 vs 2.65 mg/L; white blood cells: 6.79 vs 6.88×10⁹/L). These differences suggest that excluded participants faced higher baseline risk, likely pushing our estimates downward and underestimating true population risks.

UK Biobank is largely European-ancestry, limiting generalizability to populations with different genetics and smoking patterns. Exposure was measured at baseline only, missing relapses and reductions over time; sustained cessation benefits may therefore be underestimated. Although E-values suggest resistance to confounding, unmeasured factors could still explain part of the association. These include childhood adversity, polygenic risk, and concurrent substance use.

Future work should test these findings with repeated smoking assessments to capture relapse and dose reduction over time. Polygenic risk scores could identify gene-environment interactions and individuals who benefit most from intensive interventions. Mechanistic studies need to map the non-inflammatory pathways linking tobacco to mood changes, including HPA-axis dysfunction, nicotinic-receptor binding, cerebrovascular function, and gut microbiome alterations. Randomised trials should compare integrated cessation programs (such as varenicline plus cognitive behavioural therapy) against usual care to establish real-world effectiveness and cost-efficiency. All studies should include ethnically diverse and socioeconomically disadvantaged groups to ensure gains reach those with the highest combined burden.

## Conclusions

Smoking accounts for a sizable, preventable share of depression. The effect rises with dose and is only partly inflammatory. Quitting brings quick and lasting benefits. The drop is measurable within one year and builds over decades. Its population impact rivals many established psychiatric interventions.

Integrating evidence-based cessation into routine mental-health care could prevent approximately one in eight new cases. This approach should combine medication and behavioural counselling, with adjunct anti-inflammatory interventions for individuals with elevated inflammatory markers. This will require investment in clinician training, reimbursement, and equitable delivery, especially for socioeconomically disadvantaged and physically inactive groups. These findings support making tobacco cessation a standard part of depression prevention and care pathways.

## Data Availability

UK Biobank data are available to bona fide researchers upon application to UK Biobank (www.ukbiobank.ac.uk). Access to the de-identified dataset used in this study was granted under the UK Biobank Material Transfer Agreement (MTA) and data-use policies for Application 911377.

## List of Abbreviations

BMI: Body mass index
CI: Confidence interval
CRP: C-reactive protein
GWAS: Genome-wide association study
HbA1c: Glycated haemoglobin
HDL: High-density lipoprotein
HPA: Hypothalamic-pituitary-adrenal
HR: Hazard ratio
ICD: International Classification of Diseases
IGF-1: Insulin-like growth factor 1
IL-6: Interleukin-6
IQR: Interquartile range
LDL: Low-density lipoprotein
nAChR: Nicotinic acetylcholine receptor
NAc: Nucleus accumbens
NHS: National Health Service
PAF: Population attributable fraction
REC: Research Ethics Committee
SD: Standard deviation
SHBG: Sex hormone-binding globulin
UK: United Kingdom
VTA: Ventral tegmental area
WBC: White blood cell

## Declarations

### Ethics approval and consent to participate

UK Biobank participants provided written informed consent. UK Biobank holds NHS Research Ethics Committee approval (REC 11/NW/0382). These analyses were conducted under UK Biobank Application 911377.

### Consent for publication

Not applicable. No identifiable personal data (including images, videos, or any personally identifying information) are presented.

### Availability of data and materials

UK Biobank data are available to bona fide researchers upon application to UK Biobank (www.ukbiobank.ac.uk). Access to the de-identified dataset used in this study was granted under the UK Biobank Material Transfer Agreement (MTA) and data-use policies for Application 911377. Analysis code and scripts (R 4.5.1) will be made available at the project repository/DOI upon acceptance; a public link can be provided on request.

### Authors’ contributions

ZZ conceptualized the study. ZZ, TS, and XW performed data curation. ZZ and TS conducted formal analysis. ZZ wrote the original draft. TS, QY, and ZW contributed to writing review and editing. ZZ and JL performed visualization. BY, JS, and AY supervised the study. All authors read and approved the final manuscript.

## Acknowledgements

We thank the participants and coordinators of the UK Biobank. We acknowledge the Medical Science Data Center of Fudan University for data management support. The computations in this research were performed using the CFFF platform of Fudan University. We are grateful to colleagues for helpful discussions on study design, data curation, and analytical reproducibility.

## Additional files

### Additional file 1

**Figure S1.**
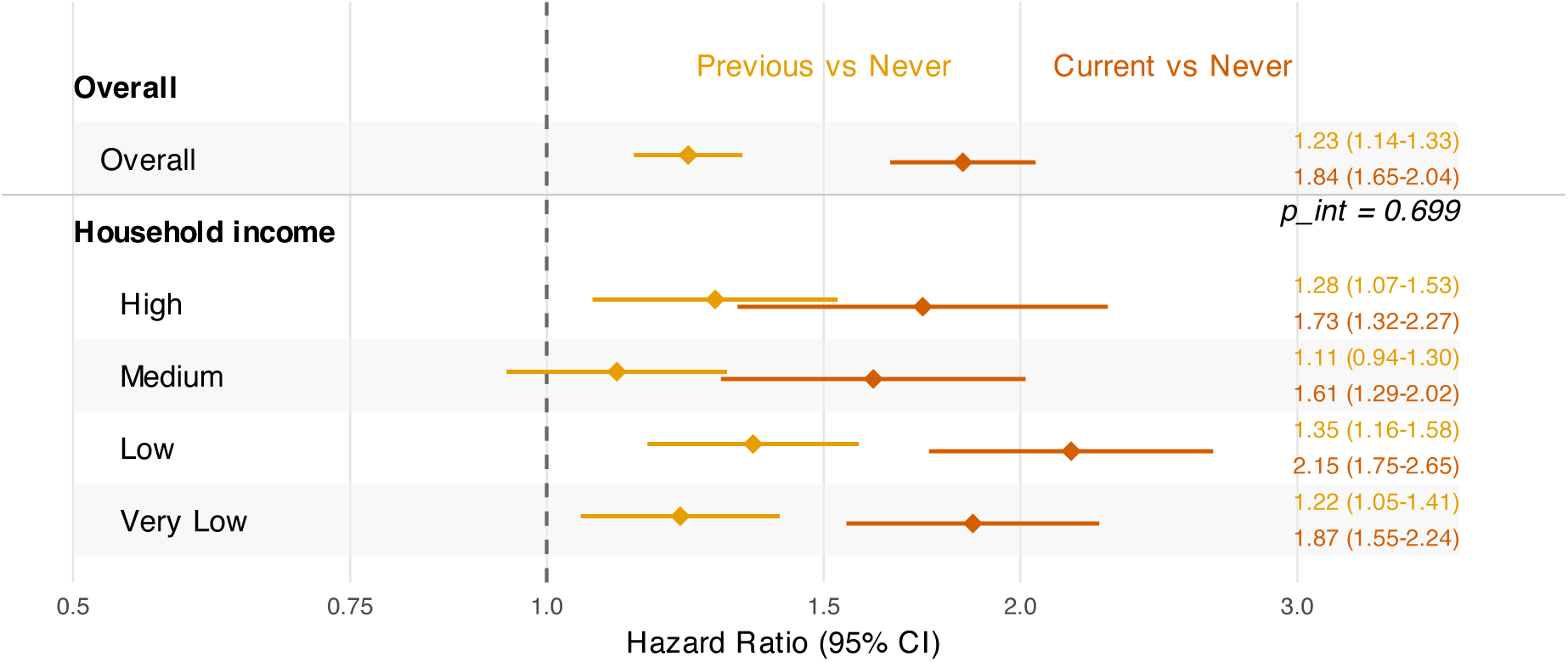
Smoking-depression associations stratified by household income. Forest plot showing hazard ratios (95% CI) for current and previous versus never smoking across four household income categories (very low, low, medium, high). Cox proportional hazards models adjusted for age, sex, socioeconomic status (excluding income), lifestyle factors, and biomarkers. P-values for interaction between smoking status and income subgroup are shown. The consistent associations across income strata show that the smoking-depression relationship holds across socioeconomic levels.

### Additional file 2

**Figure S2.**
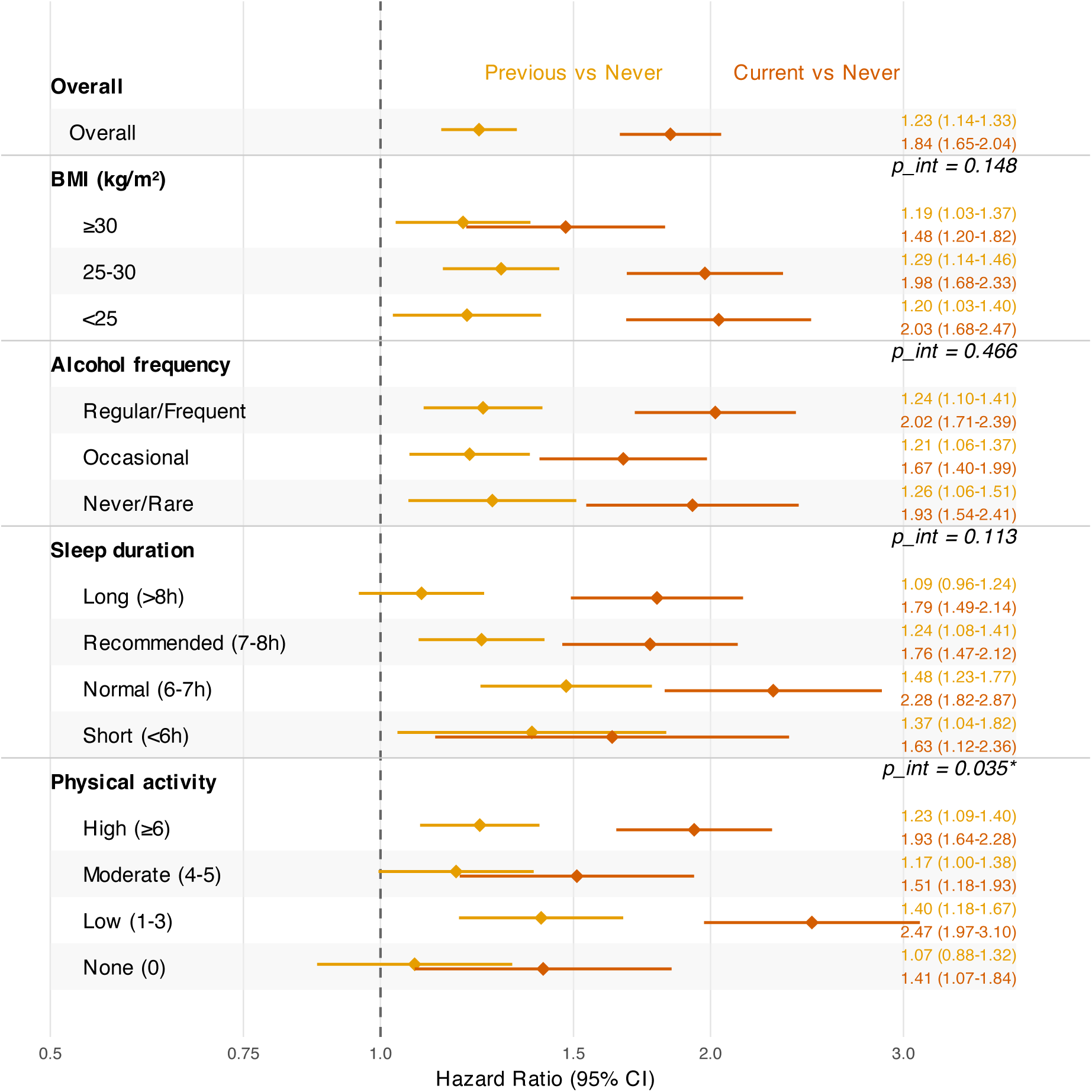
Smoking-depression associations stratified by lifestyle factors. Forest plot showing hazard ratios (95% CI) for current and previous versus never smoking across four lifestyle subgroups: BMI categories (underweight, normal, overweight, obese), alcohol frequency (never, occasional, regular), sleep duration (short, normal, recommended, long), and physical activity levels (none, low, moderate, high). Cox models adjusted for all covariates except the stratification variable. P-values for interaction terms are provided. Results demonstrate consistent smoking-depression associations across lifestyle profiles, with significant interaction for physical activity (P<0.05).

### Additional file 3

**Figure S3.**
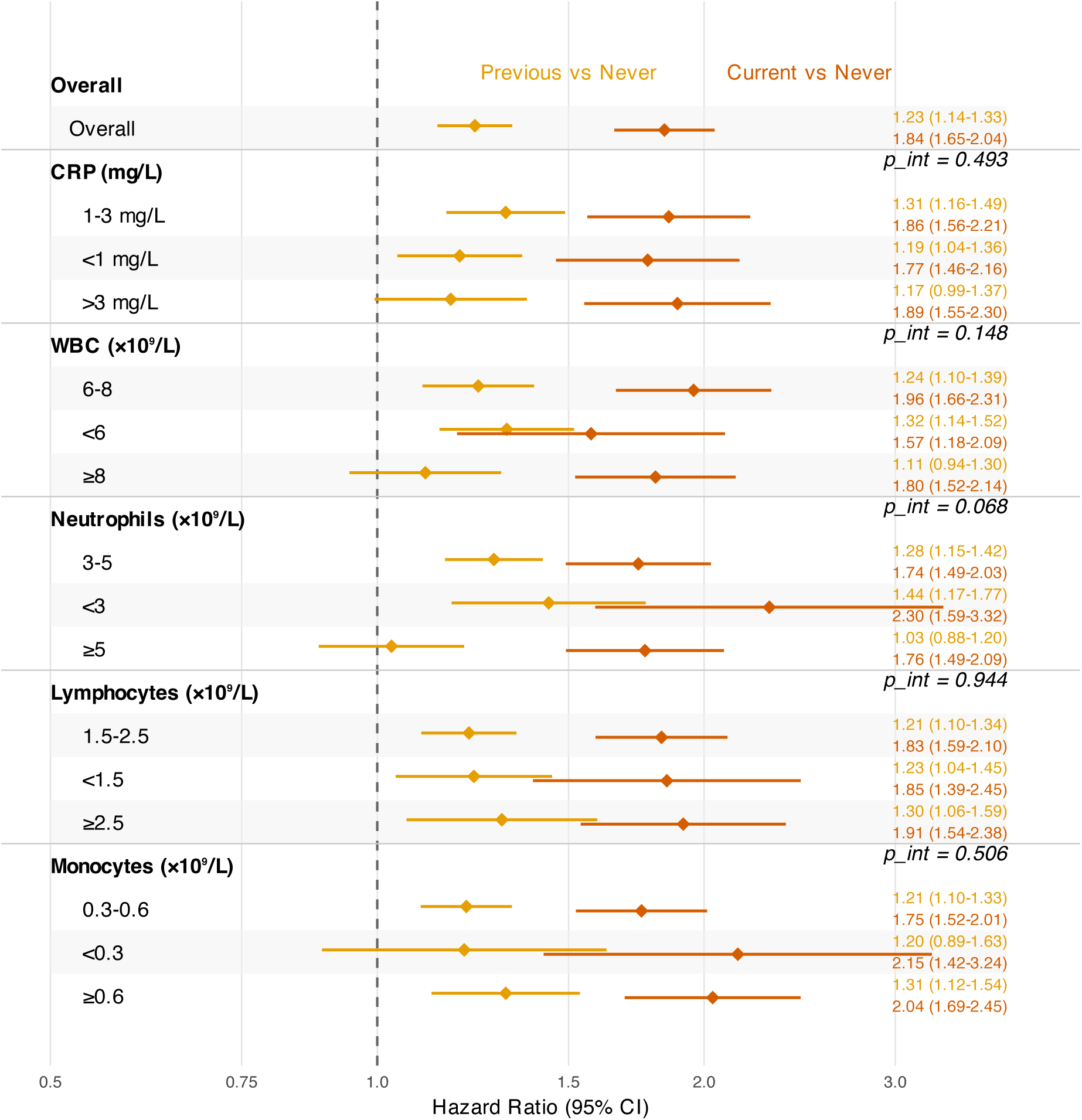
Smoking-depression associations stratified by inflammatory biomarkers. Forest plot showing hazard ratios (95% CI) for current and previous versus never smoking across tertiles or clinically defined categories of five inflammatory markers: C-reactive protein (<1, 1-3, >3 mg/L), white blood cell count (<6, 6-8, ≥8 ×10⁹/L), neutrophils (<3, 3-5, ≥5 ×10⁹/L), lymphocytes (<1.5, 1.5-2.5, ≥2.5 ×10⁹/L), and monocytes (<0.3, 0.3-0.6, ≥0.6 ×10⁹/L). Models adjusted for all covariates except the stratification biomarker. The consistent smoking effects across inflammatory strata support that smoking influences depression through both inflammatory and non-inflammatory pathways.

### Additional file 4

**Figure S4.**
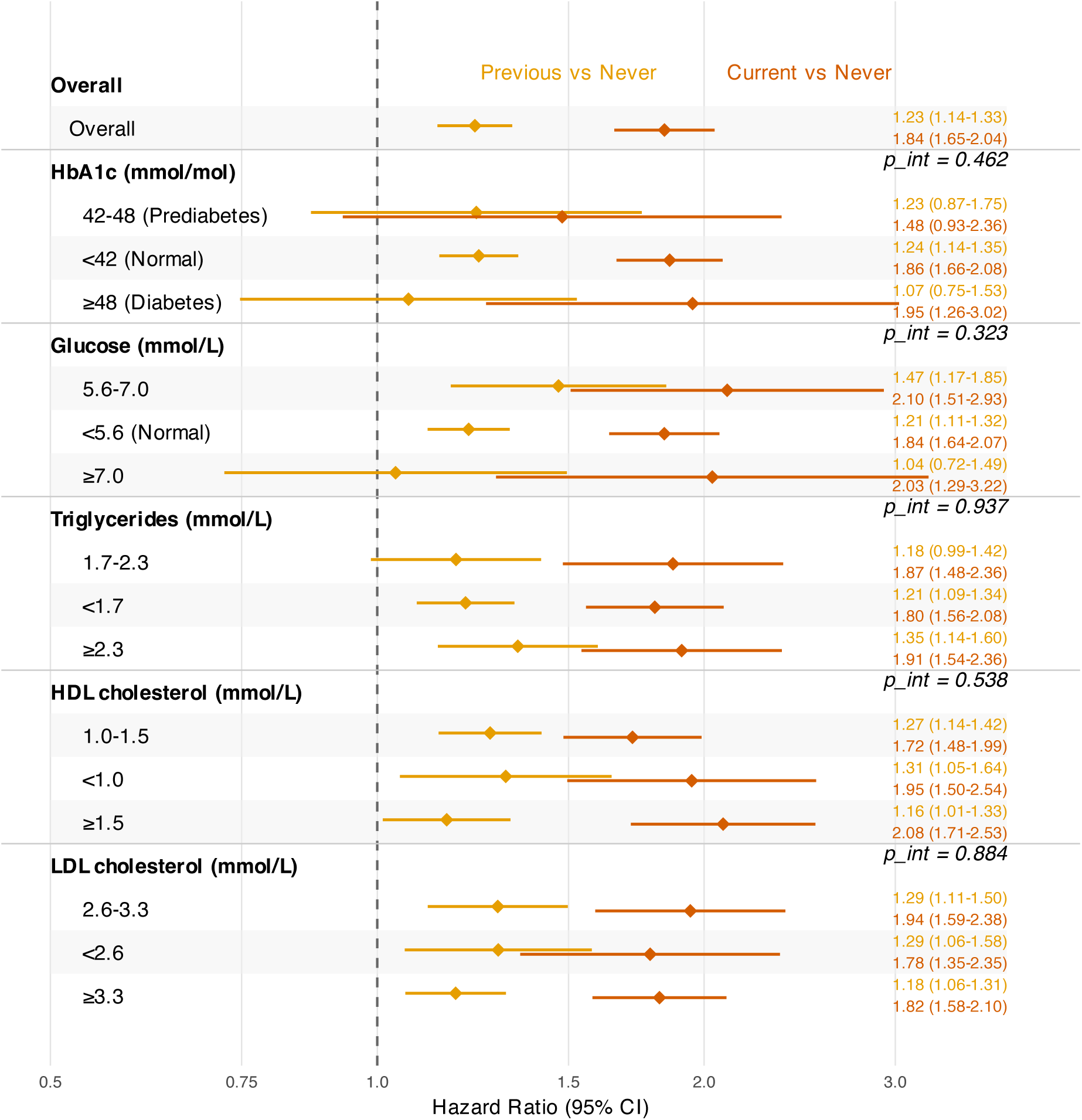
Smoking-depression associations stratified by metabolic markers. Forest plot showing hazard ratios (95% CI) for current and previous versus never smoking across clinically defined categories of five metabolic biomarkers: HbA1c (<42, 42-48, ≥48 mmol/mol corresponding to normal, prediabetes, diabetes), glucose (<5.6, 5.6-7.0, ≥7.0 mmol/L), triglycerides (<1.7, 1.7-2.3, ≥2.3 mmol/L), HDL cholesterol (<1.0, 1.0-1.5, ≥1.5 mmol/L), and LDL cholesterol (<2.6, 2.6-3.3, ≥3.3 mmol/L). Models adjusted for all covariates except the stratification marker. P-values for interaction terms are shown. Consistent associations indicate metabolic status does not substantially modify smoking-depression relationships.

### Additional file 5

**Figure S5.**
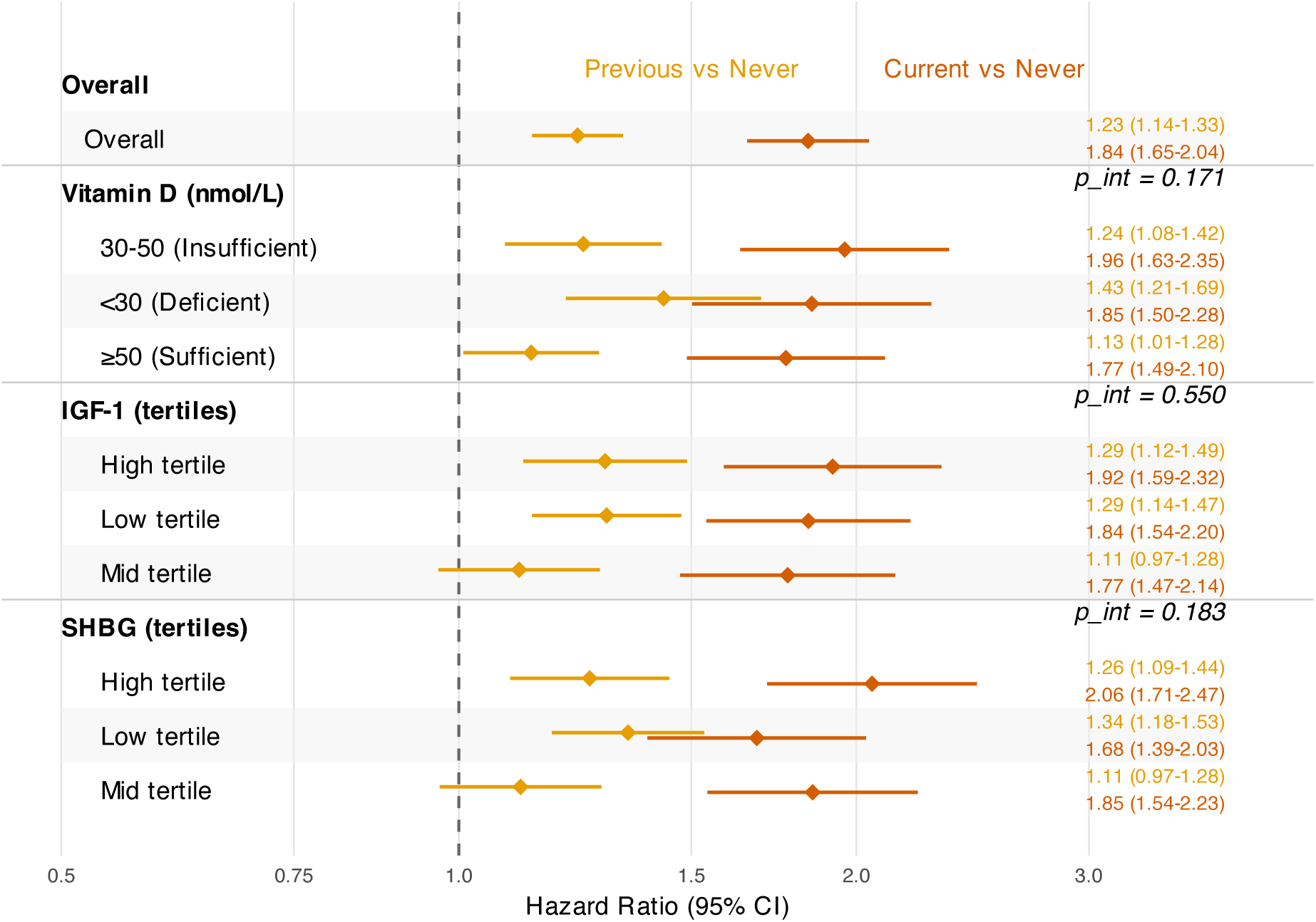
Smoking-depression associations stratified by hormonal indicators. Forest plot showing hazard ratios (95% CI) for current and previous versus never smoking across categories of three hormonal markers: vitamin D deficiency status (<30, 30-50, ≥50 nmol/L corresponding to deficient, insufficient, sufficient), insulin-like growth factor-1 (IGF-1) tertiles, and sex hormone-binding globulin (SHBG) tertiles. Cox models adjusted for all covariates except the stratification hormone. The relatively uniform associations across hormonal strata suggest that endocrine pathways do not strongly modify the smoking-depression relationship at these baseline biomarker levels.

## References

1. Reitsma MB, Kendrick PJ, Ababneh E, Abbafati C, Abbasi-Kangevari M, Abdoli A, et al. Spatial, temporal, and demographic patterns in prevalence of smoking tobacco use and attributable disease burden in 204 countries and territories, 1990–2019: A systematic analysis from the global burden of disease study 2019. Lancet. 2021;397:2337–60. 10.1016/S0140-6736(21)01169-7.

2. Global, regional, and national burden of 12 mental disorders in 204 countries and territories, 1990–2019: A systematic analysis for the global burden of disease study 2019. The Lancet Psychiatry. 2022;9:137–50. 10.1016/S2215-0366(21)00395-3.

3. Zhao Y, Yang L, Sahakian BJ, Langley C, Zhang W, Kuo K, et al. The brain structure, immunometabolic and genetic mechanisms underlying the association between lifestyle and depression. Nature Mental Health. 2023;1:736–50. 10.1038/s44220-023-00120-1.

4. Goodwin RD, Davidson L. Prevalence of cigarette smoking among US adults with major depression or substance use disorders. JAMA. 2022;328:585. 10.1001/jama.2022.10852.

5. Völker M, Streit F, Callies C, Reinhard I, Witt SH. Temporal and dose-response relationships between smoking and depression: Insights from the german national cohort (NAKO). Neuroscience Applied. 2025;4:105446. 10.1016/j.nsa.2025.105446.

6. Wootton RE, Richmond RC, Stuijfzand BG, Lawn RB, Sallis HM, Taylor GMJ, et al. Evidence for causal effects of lifetime smoking on risk for depression and schizophrenia: A mendelian randomisation study. Psychological Medicine. 2020;50:2435–43. 10.1017/S0033291719002678.

7. Pasman JA, Bergstedt J, Harder A, Gong T, Xiong Y, Hägg S, et al. An encompassing mendelian randomization study of the causes and consequences of major depressive disorder. Nature Mental Health. 2025;3:1002–11. 10.1038/s44220-025-00471-x.

8. Carroll AJ. Elucidating directionality between smoking and depression. Journal of Psychosomatic Research. 2019;125:109790. 10.1016/j.jpsychores.2019.109790.

9. Andersen AJ, Mary-Krause M, Melchior M. Smoking status trajectories, intergenerational socioeconomic mobility and depression: Preliminary results from 107,734 french adults (18 to 75 years) of the CONSTANCES cohort. European Psychiatry. 2023;66:S532–2. 10.1192/j.eurpsy.2023.1126.

10. Kim SH, Kim HC, Kim HS. Gender differences in risk of developing depression according to smoking, alcohol consumption and age: A nationwide cohort study in South Korea. Neuroscience Applied. 2023;2:102909. 10.1016/j.nsa.2023.102909.

11. Trigg J, Calabro R, Anastassiadis P, Bowden J, Bonevski B. Association of anxiety and depression symptoms with perceived health risk of nicotine vaping products for smoking cessation. Frontiers in Psychiatry. 2024;15:1277781. 10.3389/fpsyt.2024.1277781.

12. Zhao X, Davey G, Wan X. Interplay of depression, smoking intention, and smoking behavior in chinese dai adolescents. Journal of Addictions Nursing. 2023;34:211–5. 10.1097/JAN.0000000000000530.

13. Obisesan OH, Mirbolouk M, Osei AD, Orimoloye OA, Uddin SMI, Dzaye O, et al. Association between e-cigarette use and depression in the behavioral risk factor surveillance system, 2016-2017. JAMA Network Open. 2019;2:e1916800. 10.1001/jamanetworkopen.2019.16800.

14. Yang X, Evans RW, George CJ, Matthews KA, Kovacs M. Adiposity and smoking mediate the relationship between depression history and inflammation among young adults. International Journal of Behavioral Medicine. 2022;29:787–95. 10.1007/s12529-022-10060-2.

15. Zeng Y, Chourpiliadis C, Hammar N, Seitz C, Valdimarsdóttir UA, Fang F, et al. Inflammatory Biomarkers and Risk of Psychiatric Disorders. JAMA Psychiatry. 2024;81:1118–29. 10.1001/jamapsychiatry.2024.2185.

16. Mac Giollabhui N, Slaney C, Hemani G, Foley ÉM, van der Most PJ, Nolte IM, et al. Role of inflammation in depressive and anxiety disorders, affect, and cognition: Genetic and non-genetic findings in the lifelines cohort study. Translational Psychiatry. 2025;15:164. 10.1038/s41398-025-03372-w.

17. Mansournia MA, Altman DG. Population attributable fraction. BMJ. 2018;360:k757. 10.1136/bmj.k757.

18. Sudlow C, Gallacher J, Allen N, Beral V, Burton P, Danesh J, et al. UK biobank: An open access resource for identifying the causes of a wide range of complex diseases of middle and old age. PLoS Medicine. 2015;12:e1001779. 10.1371/journal.pmed.1001779.

19. Taylor G, McNeill A, Girling A, Farley A, Lindson-Hawley N, Aveyard P. Change in mental health after smoking cessation: Systematic review and meta-analysis. BMJ. 2014;348 feb13 1:g1151–1. 10.1136/bmj.g1151.

20. Lindson N, Theodoulou A, Ordóñez-Mena JM, Fanshawe TR, Sutton AJ, Livingstone-Banks J, et al. Pharmacological and electronic cigarette interventions for smoking cessation in adults: Component network meta-analyses. Cochrane Database of Systematic Reviews. 2023. 10.1002/14651858.CD015226.pub2.

21. Changeux J-P. Nicotine addiction and nicotinic receptors: Lessons from genetically modified mice. Nature Reviews Neuroscience. 2010;11:389–401. 10.1038/nrn2849.

22. Koob GF, Volkow ND. Neurobiology of addiction: A neurocircuitry analysis. The Lancet Psychiatry. 2016;3:760–73. 10.1016/S2215-0366(16)00104-8.

23. Le Borgne T, Nguyen C, Vicq E, Jehl J, Solié C, Guyon N, et al. Nicotine engages a VTA-NAc feedback loop to inhibit amygdala-projecting dopamine neurons and induce anxiety-like behaviors. Nature Communications. 2025;16:6196. 10.1038/s41467-025-61180-8.

24. Kim B, Han K, Chung H, Kim SG, Cho S-J. Lower risk of depression after smoking cessation and alcohol abstinence in patients with gastric cancer who underwent gastrectomy: A population-based nationwide cohort study. Journal of Clinical Oncology. 2023;41 4_suppl:314–4. 10.1200/JCO.2023.41.4_suppl.314.

25. Cho ER, Brill IK, Gram IT, Brown PE, Jha P. Smoking cessation and short- and longer-term mortality. NEJM Evidence. 2024;3. 10.1056/EVIDoa2300272.

26. Jawad U, Farooq R, Khan MS, Zain FU, Irfan MS, Fuaad M. Prevalence of usage nicotine and its dependence among patients with depression. Pakistan Journal of Medical and Health Sciences. 2022;16:1138–40. 10.53350/pjmhs221651138.

27. Khandaker GM, Zuber V, Rees JMB, Carvalho L, Mason AM, Foley CN, et al. Shared mechanisms between coronary heart disease and depression: Findings from a large UK general population-based cohort. Molecular Psychiatry. 2020;25:1477–86. 10.1038/s41380-019-0395-3.

28. Dai M, Wang Q. Links between psychiatric disorders and cardiovascular diseases. Phenomics. 2025;5:330–2. 10.1007/s43657-024-00217-2.

29. Han X, Zeng Y, Shang Y, Hu Y, Hou C, Yang H, et al. Risk of cardiovascular disease hospitalization after common psychiatric disorders: Analyses of disease susceptibility and progression trajectory in the UK biobank. Phenomics. 2024;4:327–38. 10.1007/s43657-023-00134-w.

30. Selvan ST, Yeo XX, Eijk Y van der. Which countries are ready for a tobacco endgame? A scoping review and cluster analysis. The Lancet Global Health. 2024;12:e1049–58. 10.1016/S2214-109X(24)00085-8.

31. Tildy B, McNeill A, East K, Gravely S, Fong GT, Cummings KM, et al. Self-reported depression and anxiety and healthcareprofessional interactions regarding smoking cessation andnicotine vaping: Findings from 2018 international TobaccoControl four country smoking and vaping (ITC 4CV) survey. Tobacco Prevention & Cessation. 2023;9 August:1–12. 10.18332/tpc/168288.

